# Longitudinal clustering of health behaviours and their association with multimorbidity: Evidence from Understanding Society (UKHLS)

**DOI:** 10.64898/2026.02.13.26346295

**Authors:** Alisha Suhag, Thomas L. Webb, John Holmes

## Abstract

**Background:** Smoking, unhealthy nutrition, alcohol consumption, and physical inactivity (SNAP behaviours) are major risk factors for multimorbidity but are often studied in isolation. Using longitudinal data, Suhag et al. identified clusters of older adults (aged ≥50) with common SNAP behaviour patterns and distinct sociodemographic profiles and multimorbidity prevalence; whether and how these patterns generalise across adulthood remains unclear.

**Aim:** To conceptually replicate Suhag et al. across a wider age range using an independent panel study.

**Methods:** We used data from Waves 7–13 of the UK Household Longitudinal Study, analysing adults (aged ≥16) participating across all seven waves (n=18,008). Repeated-measures latent class analysis identified clusters of adults with common SNAP behaviours at Waves 7, 9, 11 and 13. Multinomial and binomial logistic regression examined how clusters were associated with sociodemographic characteristics and disease status (six disease groups plus multimorbidity), respectively.

**Findings:** Seven clusters were identified: Overall Low-risk (20% of the sample), Insufficiently active (18%), Poor diet and Insufficiently active (23%), Hazardous and Harmful drinkers (11%), Hazardous drinkers, Insufficiently active and Poor diet (14%), Smokers and Drinkers (5%), and Smokers (9%). Behavioural profiles within clusters were largely stable over time. Associations between clusters and disease outcomes were counterintuitive. The cluster labelled Overall Low-risk on the basis of SNAP behaviours had the highest prevalence of multimorbidity, whereas the Hazardous drinkers, Insufficiently active and Poor diet cluster showed lower prevalence across most conditions. These clusters also differed in sociodemographic composition: the Overall Low-risk cluster comprised mainly older women with lower education and income, while the Hazardous drinkers, Insufficiently active and Poor *diet* cluster was more likely to comprise individuals in the highest education and income groups.

**Conclusion:** Cluster-analytic techniques can be used to identify population subgroups with distinct behavioural and disease profiles, underscoring the need to consider risk behaviours in conjunction with sociodemographic context.

## Introduction

Multimorbidity—the presence of two or more chronic conditions— is a growing global health challenge, affecting roughly one-third of adults [1]. Compared to individuals with a single condition, those with multimorbidity face higher risks of premature mortality, frequent hospitalisations, and prolonged healthcare utilisation [2]. In the UK, an ageing population and declining mortality rates have contributed to a high prevalence of multimorbidity, with recent cohorts exhibiting the condition at younger ages than previously observed [3–5]. Although age is a major driver, the prevalence of multimorbidity is actually higher in people aged under 65, primarily because they represent a larger portion of the population [2]. Thus, multimorbidity is not merely a consequence of aging — it is a pressing public health issue that requires proactive, targeted prevention.

A growing body of evidence suggests that key health-risk behaviours—smoking, unhealthy nutrition, alcohol consumption, and physical inactivity (collectively known as SNAP behaviours)—are important risk factors for multimorbidity [6]. However, studies diverge on which SNAP behaviours matter most: some rank smoking—alone or with physical inactivity—as the most significant risk factor [7], while reports on physical inactivity range from no independent effect [7] to a 45% increased risk [8]. Similarly, one cohort study cited age, BMI, and meat intake as top predictors (with smoking significant only in men and activity, sleep, and alcohol moderately influential) [9], while another linked women’s tobacco use to an 87% higher multimorbidity risk and alcohol consumption to an 18% increase [10].

The aforementioned studies have either investigated risk behaviours individually, in pairs, or using a cumulative risk index that sums the number of risky behaviours that an individual engages in. While such methods offer insights into population-attributable risk of disease, focusing on single behaviours is insufficient to provide insights on how multiple health behaviours combine to influence disease outcomes, whereas a cumulative index fails to identify which specific combinations of behaviours carry the greatest risk. This limits our ability to capture the specific behavioural combinations that may interact and disproportionately influence health outcomes, leaving a critical gap in the evidence needed for designing preventive interventions (i.e. whom and which behaviours or combinations of behaviour to target) [9]. Thus, while much is known about individual and paired risk factors—their prevalence, demographic correlates, and disease associations—far less is understood about how behaviours cluster (i.e. occur in specific combinations within distinct subpopulations) over time, and the health risks these clusters pose. This is important as risk behaviours and their health impacts can vary significantly across demographic groups [11–14].

In this context, latent class analysis (LCA) is a valuable tool for identifying subgroups with distinct combinations of multiple risk factors [15]. LCA is a modelling technique that aims to identify homogeneous, mutually exclusive classes (or subgroups) in a population based on similar patterns of responses. By uncovering subgroups with qualitatively different behavioural profiles, LCA allows researchers to identify those with riskier patterns and inform targeted disease prevention [11, 16]. As LCA incorporates multiple behaviours into a single latent variable, it streamlines risk assessment—eliminating the need for numerous interaction terms and lowering the probability of false-positive findings. Moreover, the exploratory nature of LCA allows behavioural profiles to be identified empirically from the observed data, without making a priori assumptions about relationships between behaviours. These advantages extend to repeated measures latent class analysis (RMLCA), a longitudinal extension of LCA that remains underutilised in areas outside child development research [17, 18]. In light of the vast repositories of population health data and an ageing population characterised by high rates of multimorbidity, techniques like latent class analysis hold potential for identifying subgroups at high risk of disease.

Despite the potential of latent modelling techniques, to date, only one study has used the technique to examine how clusters of SNAP risk behaviours are associated with multimorbidity [19]. Suhag et al. [19] identified seven clusters with stable risk behaviours over time among older adults (aged 50 years or older) in England, each with distinct sociodemographic profiles and disease prevalence. While some findings were expected—such as high-risk smokers being most likely to have respiratory disorders and low-risk clusters with higher socioeconomic status having a lower prevalence of all health conditions studied—some findings were unexpected. For example, the’Abstainer but inactive’ cluster, composed predominantly of women who were physically inactive but engaged in no other risk behaviours, had the highest prevalence of multimorbidity, even higher than clusters characterised by multiple risk behaviours [19]. This counterintuitive association persisted even after adjusting for key sociodemographic factors (age, sex, education, household income, occupation), suggesting that the relationship between behaviour and health outcomes is more complex than a linear dose–response. Crucially, the study highlighted that neither the number nor the combination of risk behaviours alone could explain why some clusters had worse health outcomes than others [19].

However, since Suhag et al.’s [19] study focused on older adults, its applicability to the general population is limited. Investigating the relationships between behaviour and disease outcomes in a younger population could provide valuable insights into the aetiological pathways and progression of multimorbidity, as age plays a pivotal role in disease development through critical exposure periods and the accumulation of risks—often occurring through’chains of risk,’ where one adverse exposure leads to another [20]. Since young people’s risk patterns tend to be less stable, and early interventions offer significant lifetime health benefits [21, 22], it is especially important to examine how risk behaviours cluster over time in younger populations.

Thus, the present study conducts a conceptual replication of Suhag et al [19] using a nationally representative UK sample (i.e. individuals aged 16 years and older). In doing so, it extends the previous research across a wider age range and has the potential to expand our understanding of the relationship between longitudinal risk behaviour clusters and multimorbidity. By elucidating how SNAP behaviours cluster over time and link to multimorbidity in the general adult population in the UK, this research aims to provide actionable insights for the development of targeted prevention strategies.

### Study Objectives

Thus, the present research analyses data from a nationally-representative, longitudinal sample of adults in the UK to:

1. Explore whether and how SNAP behaviours cluster over time in adults.
2. Investigate how membership in different behavioural clusters varies by sociodemographic characteristics.
3. Examine which behavioural clusters are prospectively associated with multimorbidity over time.

## Methods

### Study design and data

We conducted a secondary analysis of data from the UK Household Longitudinal Study (UKHLS), also known as Understanding Society—a nationally-representative panel survey of approximately 40,000 UK households followed annually since 2009–2010 [23]. UKHLS collects information across a range of domains, including health, education, employment, income and social life.

Data are collected in waves, with each wave fielded over an approximately 24-month period using a combination of face-to-face and self-completion computer-assisted interviews. The study employs a complex sampling design: clustered, stratified probability samples of households in England, Scotland, and Wales, and a systematic random sample in Northern Ireland. Full details of the UKHLS sampling strategy and data collection methods are available elsewhere [23]. To mitigate the effects of attrition, UKHLS implements sample replenishment procedures that maintain the representativeness of the cohort over time [23]. Therefore, the weighted sample starting at Wave 7 (the baseline used for the present study) remains broadly representative of the UK population.

This study adhered to the STROBE guidelines [24] (Strengthening the Reporting of Observational Studies in Epidemiology; detailed in Supplementary Section 6 in S1 Appendix). The University of Essex Ethics Committee has approved data collection for the UKHLS data, and anonymised data are available to researchers [25]. The authors accessed the data on 30 November 2023 and did not have access to any information that could identify individual participants during or after data access.

### Sample Selection and Exclusion Criteria

The analysis used data from 18,318 respondents aged 16+ who completed the main adult questionnaire across all seven waves between Wave 7 (2015–2017) and Wave 13 (2021–2023). Wave 7 was selected as the baseline because prior waves used inconsistent measures for three of the four SNAP risk behaviours (excluding smoking). Data from all waves was required to allow modelling of behavioural clusters over time.

Participants with missing sociodemographic data at baseline (n=310) were excluded, resulting in a final sample of 18,008 (98.3% of the original sample; see Figure 1). We used listwise deletion as the modelling software used for latent class regression does not support the inclusion of cases with missing data on predictor variables. As exclusions were few (∼1.7% of the original sample) and sociodemographic missingness was low (<2%), a complete-case analysis was used, as previous methodological work suggests limited risk of selection bias in such conditions [26].

**Figure 1.**
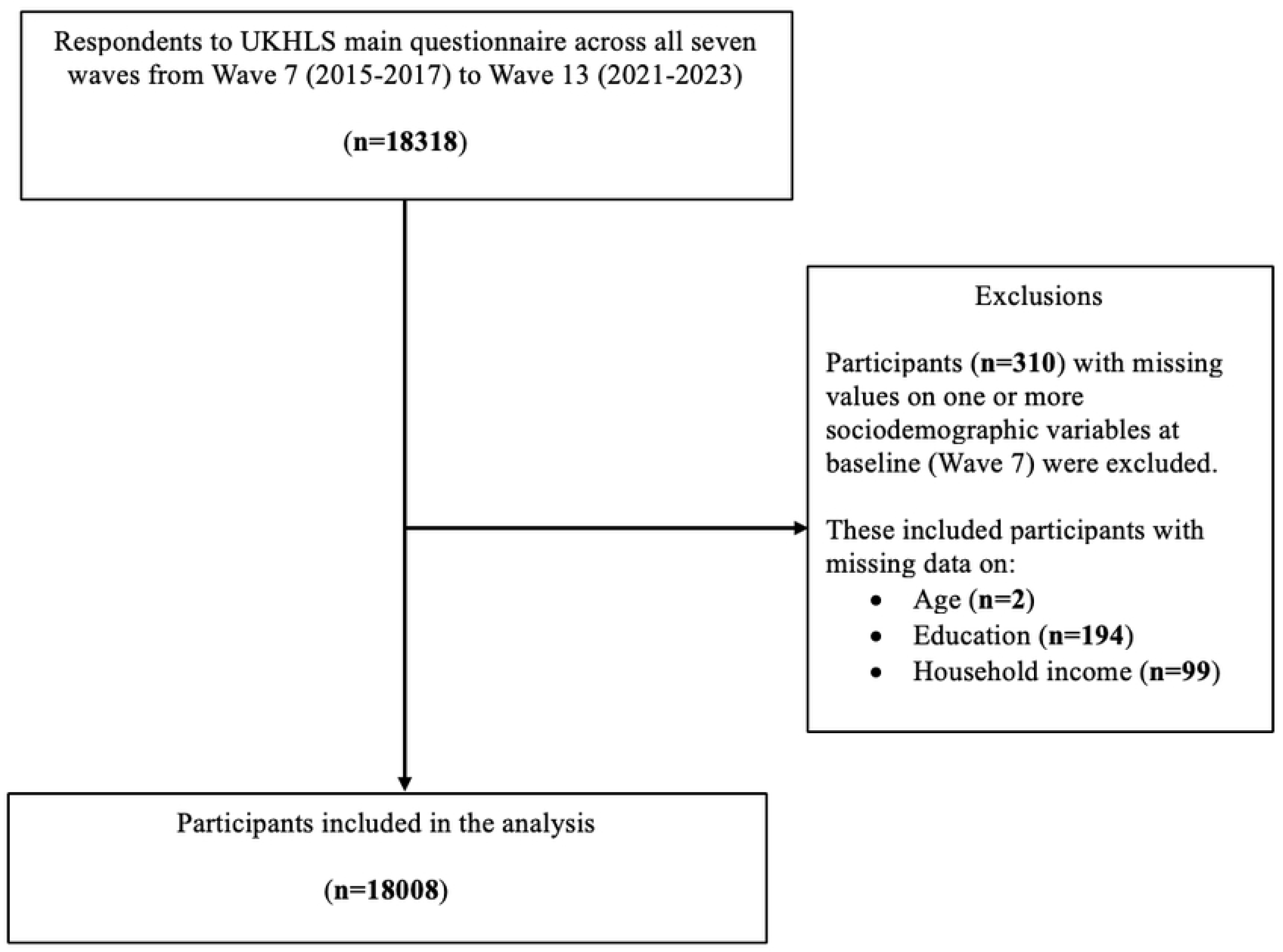
Study flow chart

All analyses utilised longitudinal survey weights provided by the UKHLS to account for non-response and sampling biases, ensuring that our results are nationally representative [23].

### Health behaviour measures

Data on the four SNAP risk behaviours were obtained from Waves 7, 9, 11, and 13 of the UKHLS. Information on smoking, fruit and vegetable consumption, and physical activity participation was collected via the main Computer-Assisted Personal Interview (CAPI), while data on alcohol consumption were gathered through a separate confidential self-completion questionnaire.

To categorise responses to the four risk behaviours, we used government recommendations or contemporary public health advice to classify levels of risky engagement (see Table 1). Further details on how engagement in these health behaviours were calculated and categorised are provided in Supplementary Section 1 in S1 Appendix.

**Table 1.** Classification of self-reported SNAP risk behaviours and their categories.

| Behaviour | Self-reported measure | Categories |
| --- | --- | --- |
| Smoking | Binary response to "Do you smoke cigarettes?" | Yes/No |
| Nutrition - Fruit and Vegetable intake | Composite of daily intake of fruit and vegetable portions | ≥5 portions/day |
|  |  | <5 portions/day |
| Alcohol consumption | AUDIT-C* score (0-12) based on frequency, quantity, and frequency of heavy episodic drinking | Low risk (0-4) |
|  |  | Hazardous drinking (5-7) |
|  |  | Harmful drinking (8-10) |
|  |  | Possible dependence (>10) |
| Physical Activity | Total MET<sup>†</sup>-min/week based on IPAQ<sup>‡</sup> short version | Low (<600 MET-min/week) |
|  |  | Moderate (600-3000 MET-min/week) |
|  |  | High (>3000 MET-min/week) |

\*AUDIT-C:Alcohol Use Disorders Identification Test-Consumption [27]
<sup>†</sup>MET: Metabolic Equivalent of Task
<sup>‡</sup>IPAQ: International Physical Activity Questionnaire [28]

### Sociodemographic variables

Data on sociodemographic variables were obtained from the baseline Wave 7 dataset, including age, sex, ethnic group, educational attainment, and monthly household income.

*Age* was grouped into 10-year bands with lower and upper bands of 16–24 years and 65 years and above.

*Ethnic group* was derived from 18 categories that participants self-identified as in the UKHLS [25], which we collapsed into: White British; White Other (Irish, Gypsy or Irish Traveller, any other White); Indian; Pakistani or Bangladeshi; Black (Black Caribbean/Black African); and Other (all other ethnicities, including mixed). Although previous studies grouped Pakistani, Bangladeshi, and Indian participants as’South Asian’ for consistency, we separated Indians due to distinct differences in alcohol abstention rates [29] and alcohol-related harms [30]. This collapsed categorisation ensured sufficient sample size in each ethnic group.

*Educational attainment* was derived from the highest level achieved and categorised as: degree or higher; A-levels or equivalent; O-levels or equivalent; other qualifications; or none.

*Household income* quintiles were derived using the UKHLS net household income variable for the month preceding the interview (including earnings, investments, and welfare benefits net of taxes and mandatory contributions), which includes imputed values. This measure was adjusted using the modified Organisation for Economic Co-operation and Development (OECD) equivalence scale to account for household size and composition. All sociodemographic variables were converted into dummy variables.

### Measures of health outcomes

Disease data were obtained from Wave 7 (baseline) and Wave 13 (final). Disease status was based on respondents’ self-reports of whether a medical doctor had ever diagnosed them with any of 16 chronic conditions and whether they still had them (see Table 2, first column). To control for baseline diseases at Wave 7, we included only health conditions assessed in both Wave 7 and Wave 13. Conditions not assessed in either wave (e.g., clinical depression, HIV, hyperthyroidism) were excluded.

**Table 2.** Chronic health conditions used to ascertain multimorbidity and their groupings.

|  | Chronic health conditions | Grouping |
| --- | --- | --- |
| 1 | Asthma | Respiratory |
| 2 | Emphysema |  |
| 3 | Chronic bronchitis |  |
| 4 | Congestive heart failure | Cardiovascular |
| 5 | Coronary heart disease |  |
| 6 | Angina |  |
| 7 | Heart attack or myocardial infarctions |  |
| 8 | Stroke |  |
| 9 | a) Bowel/colorectal cancer* | Cancers |
|  | b) Lung cancer* |  |
|  | c) Breast cancer (females only)* |  |
|  | d) Prostate cancer (males only) * |  |
|  | e) Liver cancer* |  |
|  | f) Skin cancer or melanoma* |  |
|  | g) Other cancer* |  |
| 10 | High blood pressure/hypertension | Obesity related |
| 11 | a) Diabetes† |  |
|  | b) Gestational diabetes (females during pregnancy) † |  |
|  | c) Other diabetes† |  |
| 12 | a) Osteoarthritis‡ | Arthritis |
|  | b) Rheumatoid arthritis‡ |  |
|  | c) Other arthritis‡ |  |
| 13 | Hypothyroidism or an under-active thyroid | Other chronic conditions |
| 14 | Any kind of liver condition |  |
| 15 | Multiple Sclerosis |  |
| 16 | Other longstanding/chronic condition |  |

\* Information on types of Cancer was only recorded in Wave 13, not in Wave 7
† Information on types of Diabetes types was only recorded in Wave 13, not in Wave 7
‡ Information on types of Arthritis was only recorded in Wave 13, not in Wave 7

There is no uniform approach for measuring multimorbidity across the literature [31]. We therefore adopted the widely accepted definition of basic multimorbidity as having two or more chronic conditions [32]. Respondents were coded as “yes” for multimorbidity if they had two or more conditions from the 16-condition list, and “no” otherwise. Further, since some conditions tend to co-occur, we grouped conditions into six broader categories (see Table 2, second column), primarily derived from Knies and Kumari (33). However, we adjusted their grouping by placing hypothyroidism under “other chronic conditions” instead of “autoimmunity,” since information on Type 1 diabetes was not available at the baseline wave.

In UKHLS, the measurement of chronic health conditions differed between Wave 7 and Wave 13, reflecting changes in questionnaire format and disease ascertainment procedures introduced from Wave 10 onwards, as well as differences between new and continuing respondents. To ensure comparability over time, we harmonised disease indicators by reconstructing current disease status using all available retrospective and prospective reports of diagnosis and persistence, applying wave-specific rules where necessary (for a detailed summary, see Supplementary Section 2 in S1 Appendix). Additionally, while some conditions (e.g., cancer, diabetes, and arthritis) were disaggregated into more detailed subcategories from Wave 10 onwards, we retained the original aggregated categories used in Wave 7 to ensure comparability over time.

Finally, morbid obesity was included as a separate health outcome, measured as a binary variable in UKHLS (BMI ≥ 40).

## Statistical Analysis

RMLCA was used to examine whether there were distinct classes of respondents who had similar patterns of SNAP behaviours over time. Analyses were conducted using MPlus v8.5 software and R version v4.0.3 [34, 35]. A two-stage approach was used.

In the first stage, to determine the optimal number of latent classes (Aim 1), we entered data on SNAP behaviours at each of the four waves as independent data points. We then created a series of LCA models with increasing numbers of latent classes (i.e. clusters) until the model stopped converging. Model fit was evaluated using several indices: Consistent Akaike’s Information Criterion (CAIC), Bayesian Information Criterion (BIC), adjusted Bayesian Information Criterion (aBIC), Approximate Weight of Evidence Criterion (AWE), and Vuong-Lo-Mendell-Rubin Likelihood Ratio Test (VLMR-LRT). For the likelihood-based tests such as the VLMR-LRT test, a p-value < 0.05 indicates that the model fit has not significantly improved compared to the model with one less class. Lower values of the BIC, CAIC, aBIC, and AWE indicate a better fitting model. Entropy and the smallest average latent class posterior probability were also used to assess how well the model separated individuals into distinct latent classes, with values closer to 1 indicating clearer separation for both measures. Missing data were handled using Full Information Maximum Likelihood. Reliability of the class solution was evaluated using split-half replication sensitivity analysis (see Supplementary Section 3, S1 Appendix for details).

In the second stage, the associations between the latent classes identified in the first stage and sociodemographic characteristics and disease status were examined. This was done using the 3-step Bolck, Croon, and Hagenaars (BCH) method in which individuals are assigned to the class that they have the highest posterior probability of belonging to [36]. To examine the association between sociodemographic characteristics and class membership (Aim 2), we regressed latent classes on sociodemographic variables in a series of multinomial logistic regressions, controlling for potential inaccuracies in class assignments (i.e. classification errors) that are extracted as part of the 3-step method. To assess whether the prevalence of health conditions differed across classes (Aim 3), we regressed each health outcome on the latent classes using a binomial logistic regression that adjusted for classification errors. For each health outcome, we conducted a series of binomial logistic regressions, that included: i) an unadjusted model, ii) a model adjusted for sociodemographic variables and, iii) a model adjusted for sociodemographic variables and disease status at baseline (we did not control for morbid obesity at the baseline wave since it was only measured in Wave 13). We adopted this stepwise adjustment approach to assess how covariate adjustment affected the associations with disease outcomes and because adjusting for baseline disease tended to reduce the analytical sample (see last column of Table 7). Then, for each health outcome in each set of these models, an omnibus Wald chi-square test was conducted to test for differences in disease prevalence across all latent classes (α=0.05). If significant, pairwise comparisons were conducted to test for differences in disease prevalence between each pair of classes, at the Bonferroni corrected significance level of α = 0.002.

## Results

### Sample characteristics

The sample (n=18,008) was predominantly White British (83.6%), with a slight female majority (56.9% females), and the average age was 50.8 years (Range=16–95, SD=16.2). In total, 43.4% of the sample possessed a degree and 8.5% held no educational qualifications (see Table 3).

**Table 3.** Sociodemographic characteristics of the final sample at baseline (n=18,008)

| **Sociodemographic characteristics at Wave 7** |  | **Study sample**<br>**(n=18008)**<br>**N (%)** |
| --- | --- | --- |
| Sex | Male | 7,760 (43.1) |
|  | Female | 10,248 (56.9) |
| Ethnicity | White British | 15,050 (83.6) |
|  | White - Other | 795 (4.4) |
|  | Indian | 574 (3.2) |
|  | Pakistani and Bangladeshi | 565 (3.1) |
|  | Black (African/Caribbean) | 414 (2.3) |
|  | Other (including mixed ethnicities) | 610 (3.4) |
| Education | Degree level or higher | 7,817 (43.4) |
|  | A-levels or equivalent | 3,627 (20.1) |
|  | O-levels or equivalent | 3,502 (19.4) |
|  | Other educational qualifications | 1,525 (8.5) |
|  | None | 1,537 (8.5) |
| Household<br>income (in<br>quintiles) | 5th quintile (Highest) | 3,624 (20.1) |
|  | 4th quintile | 3,600 (20.0) |
|  | 3rd quintile | 3,601 (20.0) |
|  | 2nd quintile | 3,602 (20.0) |
|  | 1st quintile (Lowest) | 3,581 (19.9) |
| Age<br>groups | 16-24 years | 1,292 (7.2) |
|  | 25-34 years | 1,964 (10.9) |
|  | 35-44 years | 2,970 (16.5) |
|  | 45-54 years | 3,811 (21.2) |
|  | 55-64 years | 3,790 (21.0) |
|  | 65 years and older | 4,181 (23.2) |

The sample’s engagement in health risk behaviours across waves is shown in Table 4. Most participants were non-smokers (around 87–91% across waves), comparable to recent national estimates [37]. Around half of the sample did not meet the recommendation of ≥5 portions/day of fruit and vegetables [38] and about one third did not meet WHO recommendations for physical activity [39]. Approximately three quarters of participants were classified as low-risk or hazardous drinkers and around 6% as harmful or possibly dependent.

**Table 4.** SNAP behaviours among participants (n= 18008) included in the latent class analysis.

|  |  | **Wave 7**<br>**(2015-17)** | **Wave 9**<br>**(2017-19)** | **Wave 11**<br>**(2019-21)** | **Wave 13**<br>**(2021-23)** |
| --- | --- | --- | --- | --- | --- |
|  |  | N (%) | N (%) | N (%) | N (%) |
| **Smoking** | Non-smoker | 15647 (86.9) | 16039 (89.1) | 16114 (89.5) | 16321 (90.6) |
|  | Smoker | 2083 (11.6) | 1889 (10.5) | 1838 (10.2) | 1647 (9.1) |
|  | Missing | 278 (1.5) | 80 (0.4) | 56 (0.3) | 40 (0.2) |
| **Fruit and vegetable intake** | ≥ 5 portions/day | 8011 (44.5) | 9688 (53.8) | 9622 (53.4) | 9499 (52.7) |
|  | < 5 portions/day | 9997 (55.5) | 8320 (46.2) | 8386 (46.6) | 8509 (47.3) |
| **Alcohol consumption** | Low-risk | 9356 (52.0) | 9440 (52.4) | 9656 (53.6) | 9512 (52.8) |
|  | Hazardous | 3923 (21.8) | 3977 (22.1) | 3550 (19.7) | 3130 (17.4) |
|  | Harmful | 1039 (5.8) | 1051 (5.8) | 953 (5.3) | 942 (5.2) |
|  | Possible dependence | 43 (0.2) | 47 (0.3) | 47 (0.3) | 46 (0.3) |
|  | Missing | 3647 (20.3) | 3493 (19.4) | 3802 (21.1) | 4378 (24.3) |
| **Physical activity** | High | 4249 (23.6) | 6016 (33.4) | 5366 (29.8) | 5069 (28.1) |
|  | Medium | 8299 (46.1) | 7481 (41.5) | 6829 (37.9) | 7034 (39.1) |
|  | Low | 5175 (28.7) | 4412 (24.5) | 5726 (31.8) | 5789 (32.1) |
|  | Missing | 285 (1.6) | 99 (0.5) | 87 (0.5) | 116 (0.6) |

In terms of disease outcomes, at Wave 13, the most prevalent conditions were: obesity-related conditions (21.3%), multimorbidity (21.2%), other chronic conditions (19.2%), and arthritis (15.2%) (see Supplementary Table 2). Less prevalent conditions included respiratory disorders (10.5%), cardiovascular diseases (6.8%), and cancer (2.3%). These prevalence rates are similar to those reported in the broader UK population [33, 40]. In addition, the prevalence of morbid obesity (BMI ≥ 40) was 3.4% in Wave 13; corresponding data were not available for Wave 7.

### Clusters of health behaviour and within-cluster changes

The selection of the optimal latent class model was guided by examining key statistical criteria (see Table 5). The VLMR-LRT test supported both five-and seven-class solutions. These models also met the recommended thresholds for entropy (≥0.8) and smallest class size [≥ 5%; 35]. However, the values of the loglikelihood, BIC, aBIC, and CAIC continued to decrease as the number of classes increased, indicating improved model fit [41]. Since the decline in information criteria plateaued around five classes (scree plot shown in Supplementary Figure 5 in S1 Appendix), we decided to further compare the five-class and seven-class models using a sensitivity analysis on random split-halves of the data. This analysis confirmed the robustness of the 5-class and 7-class solutions, as identical classes were uncovered in each split-half (full details can be found in Supplementary Section 3 in S1 Appendix).

**Table 5.** Model fit evaluation information for choosing a latent class model.

| K | LL | CAIC | BIC | aBIC | AWE | VLMR<br>LRT<br>p-value | Entropy | Smallest<br>average<br>latent class<br>posterior<br>probability | Smallest<br>class size<br>(%) |
| --- | --- | --- | --- | --- | --- | --- | --- | --- | --- |
| 1 | -203281.5 | 406619 | 406838 | 406749 | 406879.4 | - | - | 1 | 100 |
| 2 | -189807.2 | 379728 | 380173 | 379992 | 380258.4 | <0.001 | 0.974 | 0.984 | 14.6 |
| 3 | -181641.8 | 363456 | 364126 | 363853 | 364255.3 | 0.002 | 0.876 | 0.916 | 13.9 |
| 4 | -176776.9 | 353784 | 354681 | 354315 | 354853.1 | 0.003 | 0.821 | 0.879 | 13.6 |
| 5 | -174867.4 | 350023 | 351146 | 350688 | 351361.8 | **0.004** | 0.805 | 0.798 | 13.6 |
| 6 | -173497.1 | 347340 | 348689 | 348140 | 348948.9 | 0.105 | 0.790 | 0.763 | 10.8 |
| **7** | **-172230.3** | **344865** | **346440** | **345798** | **346742.9** | **0.031** | **0.796** | **0.759** | **5.1** |
| 8 | -171261.4 | 342985 | 344786 | 344052 | 345132.8 | 0.234 | 0.777 | 0.756 | 5.1 |
| 9 | -170382.3 | 341285 | 343312 | 342486 | 343702.2 | 0.080 | 0.777 | 0.751 | 4.6 |

*Note:* $n=18008$ ; K = number of classes (the nine-class model failed to converge); LL = model
log likelihood; BIC = Bayesian information criterion; aBIC = sample size adjusted BIC; CAIC
= consistent Akaike information criterion; AWE = approximate weight of evidence criterion;
VLMR-LRT = Vuong-Lo-Mendell-Rubin adjusted likelihood ratio test; p-value significance

Given that these statistical indices did not uniformly point to a single model specification (and may over-or underestimate the number of classes present), the specific model estimates for both 5-and 7-class models were examined to select a final model based on whether it was possible to label the latent classes meaningfully, as well as theory and previous findings [19, 42, 43]. Based on these criteria, the 7-class model was deemed the most appropriate fit to the data (see Supplementary Section 4 in S1 Appendix for a detailed comparison between the 5-and 7-class models).

The seven distinct health-risk behaviour classes — hereafter referred to as clusters — were assigned labels by the research team. Figure 2 depicts these clusters and the average probability that individuals in each cluster engaged in the four health behaviours—smoking, alcohol consumption, physical activity, and fruit and vegetable intake—over four distinct time points. Most behaviours were fairly stable over time, so unless highlighted below, the behaviours that characterise each cluster were similar at each time point.

**Figure 2.**
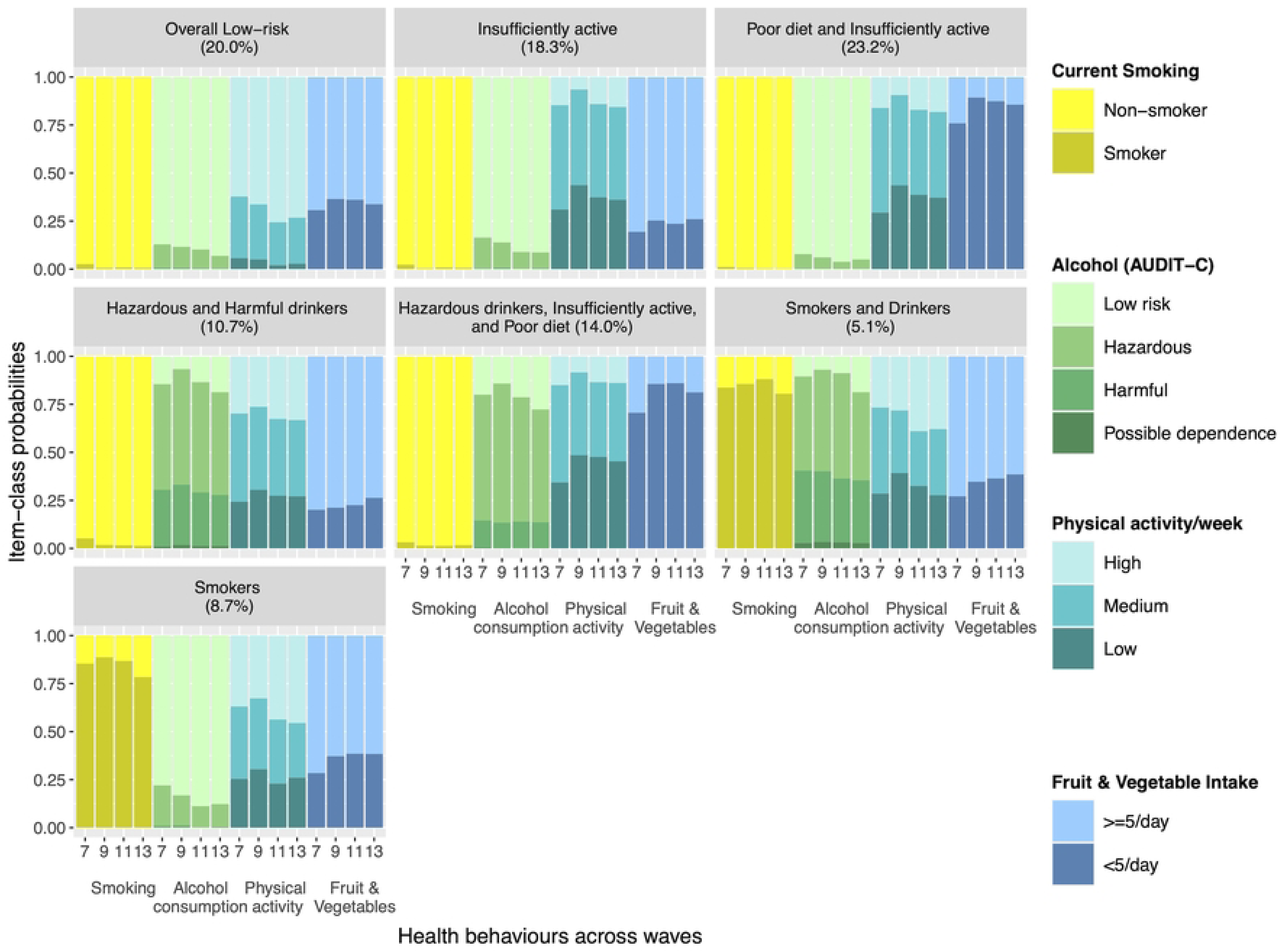
Seven-class model reflecting different clusters of health behaviour across time ***Note.*** The x-axis lists each of the four behaviours – smoking, alcohol consumption, physical activity, and fruit and vegetable intake – across waves 7, 9, 11 and 13. The y-axis provides the average probability for each of the indicators (i.e. four health behaviours) conditional on membership in a given class (i.e. cluster).

Participants in the cluster labelled *Overall Low-risk* (20% of the sample) maintained adequate (≥5 per day) fruit and vegetable intake and high levels of physical activity across time. They had a consistently high probability of drinking at low-risk levels, and the proportion of people consuming alcohol at hazardous levels fell across waves.

Participants in the cluster labelled *Insufficiently active* (18.3%) displayed low-to-medium levels of physical activity. However, they consumed the recommended portions of fruit and vegetables, and had low-risk alcohol intake.

The largest proportion of participants fell in the cluster labelled *Poor diet and Insufficiently active* (23.2%). Participants in this cluster exhibited patterns of smoking and alcohol consumption similar to those in the cluster labelled *Overall Low-risk*. However they had a high probability of having inadequate fruit and vegetable intake and low-medium levels of physical activity.

Participants in the cluster labelled *Hazardous and harmful drinkers* (10.7%) had a high likelihood of consuming alcohol at hazardous or harmful levels. They consumed the recommended portions of fruit and vegetables and had the same probability of engaging in physical activity across all three levels (i.e. high, medium, low).

Participants in the cluster labelled *Hazardous drinkers, Insufficiently active and Poor diet* (14%) had a similar profile to the cluster labelled *Poor diet and Insufficiently active,* except they had a consistently high probability of consuming alcohol at hazardous levels.

Participants characterised as *Smokers and drinkers* (5.1%) had a high probability of smoking, increasing over time then declining over the last wave, adequate fruit and vegetable intake and low levels of physical activity. Notably, participants in this cluster exhibited patterns of alcohol consumption and physical activity similar to those in the *Hazardous and harmful drinkers* cluster.

Finally, participants in the cluster labelled *Smokers* (8.7%) had a high probability of smoking, albeit declining over time, medium to high levels of physical activity, and adequate fruit and vegetable intake.

### Sociodemographic characteristics of participants in each cluster

Table 6 displays the sociodemographic composition of participants in each of the identified clusters and the results from the adjusted multinomial logistic regressions that examined associations between each sociodemographic characteristic and cluster membership.

**Table 6.**
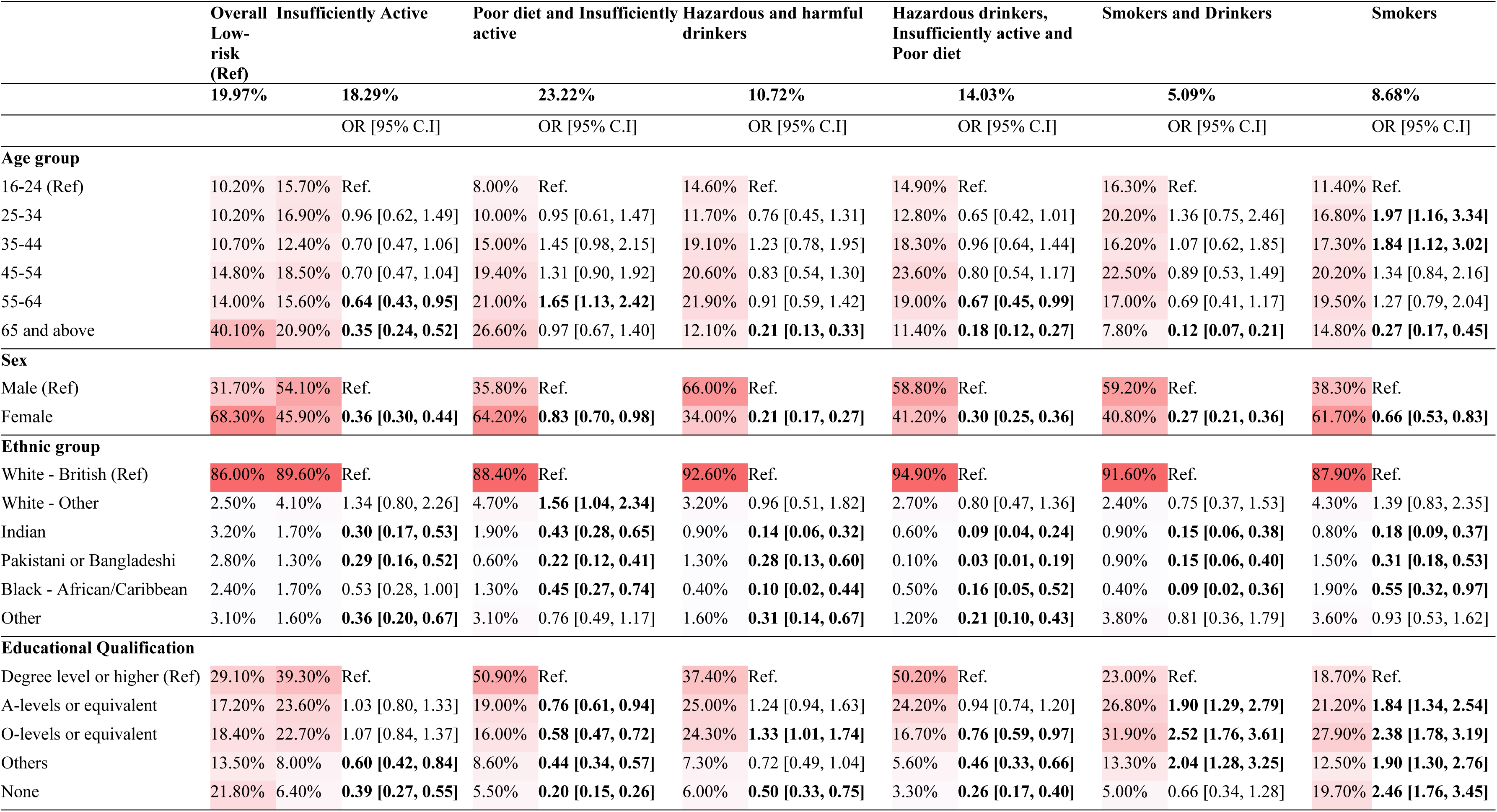
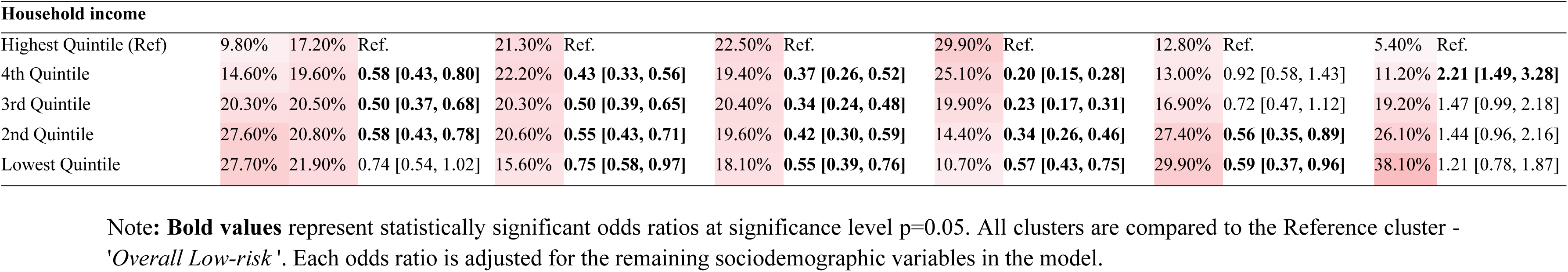
Association between sociodemographic predictors and cluster membership, adjusted for all other sociodemographic predictors.

**Table 7.**
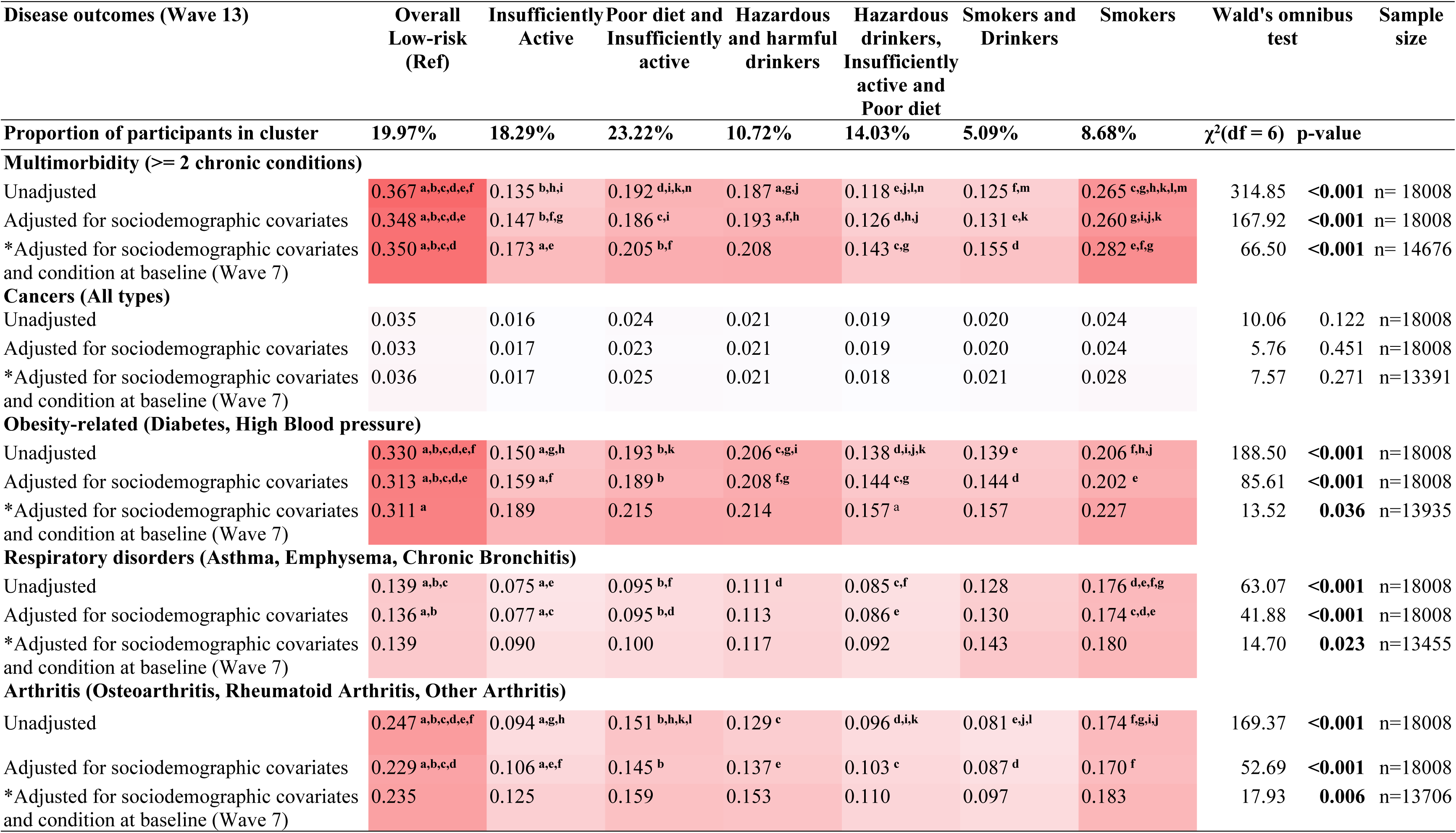
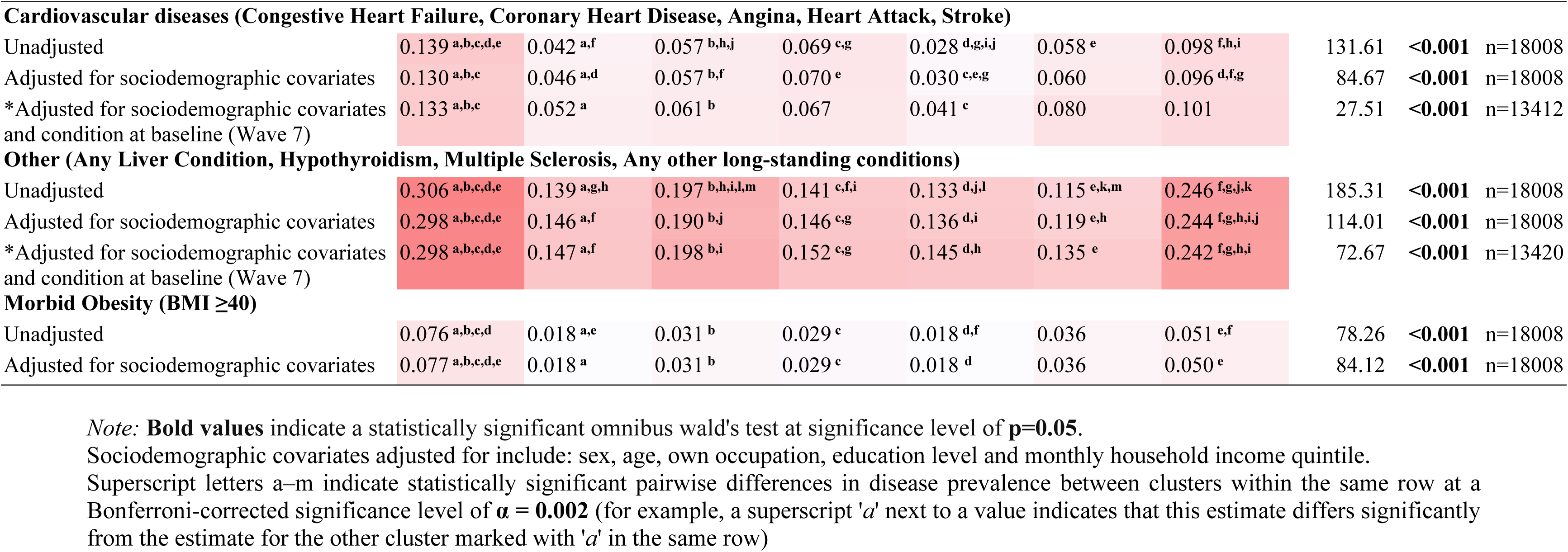
Pairwise comparisons of disease status prevalence across clusters.

Compared to participants in the *Overall Low-risk* cluster (reference group), participants characterised as *Smokers and Drinkers* and *Smokers* were about twice as likely to be aged 25-44 years than to fall in the youngest age group (reference category). Conversely, participants in the *Overall Low-risk* cluster were more likely to be 65 years and older than in the youngest age group, compared to participants in all other clusters except the *Poor diet and Insufficiently active* cluster. In turn, participants in the *Poor diet and Insufficiently active* cluster, were around 1.7 times more likely to be aged 55-64 years compared to participants in the *Overall Low-risk* cluster.

Women constituted approximately two-thirds of the participants in both the *Overall Low-risk* and *Poor diet and Insufficiently active* clusters, while men made up two-thirds of the *Hazardous and harmful drinkers.* Notably, the *Overall Low-risk* cluster had a higher likelihood of female membership compared to all other clusters.

Aligning with the ethnic distribution of UKHLS, participants in all clusters were predominantly White British (the reference category). However, participants in the *Poor diet and Insufficiently active* cluster were 1.5 times more likely to belong to other White ethnicities compared to participants in the *Overall Low-risk* cluster. In contrast, participants in *Overall Low-risk* cluster were more likely to belong to ethnic minorities such as Indian, Pakistani or Bangladeshi, or Black as well as ethnicities categorised as other (which included mixed ethnicities) compared to participants in all other clusters.

Participants in two clusters – *Poor diet and Insufficiently active* and *Hazardous drinkers, Insufficiently active and Poor diet* – had the highest proportion (approximately 50%) of individuals educated to degree level or higher. Conversely, participants in the *Smokers and Drinkers* and *Smokers* clusters were nearly twice as likely to have their highest educational qualification at other levels (i.e. qualifications below degree level), compared to participants in the *Overall Low-risk* cluster. Notably, those in the *Smokers* cluster were 2.5 more likely to have no educational qualifications compared to the *Overall Low-risk* cluster.

Compared to participants in the *Overall Low-risk* cluster, participants in three clusters—namely,’*Poor diet and Insufficiently active; Hazardous and harmful drinkers; and Hazardous drinkers*, *Insufficiently active and Poor diet*—had a higher likelihood of belonging to the highest quintile of household income, rather than any other quintiles. Conversely, participants characterised as *Smokers* were twice as likely to belong to the fourth (i.e. second highest) quintile of household income compared to participants in the *Overall Low-risk* cluster

### Relationship between health behaviour clusters and disease status

In unadjusted analyses, the prevalence of multimorbidity, respiratory disorders, arthritis, cardiovascular diseases, obesity-related conditions, morbid obesity, and conditions classified as ‘other’ differed significantly across clusters, as indicated by Wald’s omnibus tests (see Table 7). To further investigate which clusters differed in the proportions of participants with each disease profile, we conducted pairwise comparisons between clusters, shown in Table 7.

Participants characterised as *Overall Low-risk* had a markedly higher prevalence of multimorbidity, obesity-related conditions and arthritis than those in all other clusters, and also a higher prevalence of cardiovascular diseases and’other’ conditions than all other clusters except *Smokers*. Participants in the *Smokers* cluster had the highest prevalence of respiratory disorders, followed by the *Overall Low-risk* cluster. The *Insufficiently Active* cluster and *Hazardous drinkers, Insufficiently active and Poor diet* had a significantly lower prevalence of respiratory disorders, obesity-related diseases, morbid obesity and cardiovascular diseases than most clusters, with the exception of the *Smokers and Drinkers* cluster. *Smokers and drinkers* had the lowest prevalence of arthritis and’other’ chronic conditions, and the second lowest prevalence of multimorbidity. The low prevalence may be partially attributed to the fact that this cluster had the highest proportion of individuals in the youngest age groups (between 16 and 34 years). Nonetheless, participants in the clusters *Overall Low-risk* and *Smokers* tended to have higher levels of most outcomes than other clusters.

Adjusting for sociodemographic characteristics led to some attenuation of effect sizes but did not materially alter the overall pattern of results (Table 7). In particular, the higher prevalence of multimorbidity, morbid obesity, obesity-related diseases and’other’ chronic conditions in the *Overall Low-risk* cluster compared with most other clusters persisted after sociodemographic adjustment.

When we additionally adjusted for the presence of the corresponding chronic condition group at baseline (Wave 7), omnibus Wald tests remained statistically significant for all disease groups except cancer, indicating residual heterogeneity in disease burden across clusters even after accounting for baseline morbidity. The proportional ordering of clusters by disease prevalence was also largely preserved: for multimorbidity, obesity-related conditions and morbid obesity, the relative ranking of clusters changed little, and only respiratory disorders showed a change in ordering, with the *Smokers* and *Smokers and Drinkers* clusters exhibiting the highest and second-highest prevalence, respectively, after baseline disease adjustment. However, several pairwise differences—particularly for obesity-related diseases, arthritis and respiratory disorders—no longer met the Bonferroni-corrected threshold.

## Discussion

This study extends previous research examining the relationship between clusters of health-risk behaviours over time and multimorbidity in older adults [22] to a nationally representative sample of UK adults aged 16 years and over. We identified seven clusters representing distinct profiles of health behaviour. Although differences in measurement limit direct comparison, these clusters broadly resemble those reported in prior UK [19, 44, 45], Irish [46], and Australian [47] studies, including an overall low-risk cluster, a physically inactive cluster with poor diet, a cluster characterised by heavy alcohol consumption, and a smoking cluster.

Beyond identifying behavioural clusters, we examined how patterns of behaviour within each cluster change over time. Overall, behaviour patterns were largely stable over the nine-year follow-up period, consistent with findings from older adults in England [19], and broader evidence that health-risk behaviours change less in mid-and late-adulthood than in adolescence and early adulthood [21, 42, 48–50]. However, there were some exceptions to this stability: Hazardous alcohol consumption decreased over time (i.e. as participants aged) in the *Overall Low-risk* and *Poor diet and Insufficiently active* clusters, which had the highest proportion of older individuals and a two-thirds female majority. This pattern of diminishing alcohol consumption in older adults, particularly women, aligns with findings from the ELSA dataset reported by Suhag et al. [19] and broader drinking trends in the UK [51]. In contrast, the proportion of current smokers followed a rise-then-fall pattern in the *Smokers and Drinkers* cluster (the youngest cluster), but declined steadily in the *Smokers* cluster, mirroring the UK-wide decline in smoking after age 25 [52]. These exceptions suggest that when change occurs, health-risk behaviours tend to improve with age.

The clusters differed in their sociodemographic profiles. Participants in the three hazardous and harmful drinking clusters were predominantly male, consistent with UK-wide alcohol consumption patterns [11], and *Smokers and Drinkers* had the highest proportion of younger adults of any cluster, with over half aged 44 years or younger [53]. In contrast, the clusters characterised as *Overall Low-risk* on the basis of their health behaviours and *Poor diet and Insufficiently active* primarily comprised older women, with roughly half aged ≥55 years. The *Poor diet and Insufficiently active* cluster—older, socioeconomically advantaged women with relatively healthy behaviours—closely resembles the’Healthy’ or’Safe’ clusters identified in prior studies [19, 46, 47]. Although participants from ethnic minority groups comprised less than 5% of all groups, they were more likely to be in the *Overall Low-risk* cluster, aligning with prior findings of healthier behaviours reported among these groups in the UK [43, 54]. Conversely, participants in the *Overall Low-risk* and *Smokers* clusters were less likely to be in the highest income quintile, more likely to have no educational qualifications, and predominantly female. The sociodemographic profile of *Smokers* likely reflects the higher prevalence and greater harms of smoking among disadvantaged groups, with the higher proportion of women potentially indicating survivorship effects given higher smoking-related mortality in men [55, 56].

The clusters also differed in their disease status at Wave 13, even after accounting for baseline disease. Participants in the *Smokers* and *Smokers and Drinkers* clusters had the highest prevalence of respiratory disorders, consistent with established links between smoking and respiratory disease [57]. However, there were two notable and unexpected findings. First, participants in the *Overall Low-risk* cluster had the highest prevalence of multimorbidity, cardiovascular diseases, morbid obesity, obesity-related and other chronic conditions, among all clusters, despite having what is typically considered to be the least risky behaviour profile (i.e., low levels of smoking and alcohol consumption, high levels of physical activity and meeting recommended fruit and vegetable intake). Second, participants in the *Hazardous drinkers, Insufficiently active and Poor diet* cluster had a lower prevalence of respiratory disorders, obesity-related diseases, morbid obesity and cardiovascular diseases than other clusters, despite engaging in three risky behaviours.

Several explanations may account for the poorer outcomes observed in the *Overall Low-risk* cluster. One possibility relates to measurement limitations. SNAP indicators did not capture key risk factors such as diet quality, sedentary time, or psychological stress [58]. In UKHLS, diet was measured using weekly fruit and vegetable intake, which captures only part of dietary risk, as diet quality depends on both nutrient density and total energy intake [59, 60]. The *Overall Low-risk* cluster also had the highest prevalence of morbid obesity, a strong predictor of multimorbidity [61]. However, because obesity was only measured at the final wave (Wave 13), we could not assess its temporal relationship with disease onset, limiting interpretation based solely on behavioural profiles.

Residual confounding may also contribute to these findings. The *Overall Low-risk* cluster predominantly comprised older women with lower household income and education. Consistent with this, a recent study found that among women, older age, lower education, and overweight status were associated with multimorbidity, whereas unhealthy SNAP behaviours were not independent predictors [62]. Although differences in multimorbidity persisted after adjustment for sociodemographic factors in our study, such adjustment may not fully capture unmeasured differences in health status and cumulative socioeconomic disadvantage. Similar issues have been documented in the alcohol epidemiology literature, where associations persist after socioeconomic adjustment but are interpreted as artefacts of residual confounding [63]. For example, moderate drinkers appear healthier than abstainers or heavy drinkers not because of alcohol itself, but because they differ in unmeasured characteristics such as baseline health, the “sick quitter” effect, and social disadvantage that standard socioeconomic indicators do not capture [63].

The second counterintuitive finding—that participants in the *Hazardous drinkers, Insufficiently active and Poor diet* cluster had a lower prevalence of most health conditions (except’other’ conditions and arthritis) — may also be partially explained by sociodemographic composition and residual confounding. This cluster was most likely to be in the highest income quintile, highly educated, and from non-minority ethnic groups. This pattern is consistent with the vulnerability hypothesis, whereby deprived groups experience disproportionate harm from unhealthy lifestyles, not merely due to a higher prevalence of ostensibly risky behaviour [64, 65].

### Strengths and limitations

The study is the first to explore the relationship between longitudinal clusters of multiple health-risk behaviours and multimorbidity in a nationally-representative sample of UK adults aged 16 years and over [66]. It uses a robust, model-based, probabilistic approach (namely, RMLCA), that has several advantages over other traditional methods used in lifestyle risk behaviour [67, 68] in that it: i) models multiple behaviours simultaneously, ii) yields stable results, and iii) is reproducible since diagnostics are accessible and programming codes can be shared upon request.

While our study benefits from several strengths, the findings should be interpreted in light of the following limitations. First, self-report measures of chronic diseases and risk behaviours may introduce biases, although longitudinal analyses are less susceptible to misclassification because of consistent measures across survey waves. Second, some risk behaviours—particularly diet— were assessed using measures that are unlikely to fully capture important differences between participants. However, such measures are common within the epidemiological literature (e.g., assessing fruit and vegetable intake rather than overall diet quality) [69]. Third, we focused on prevalence rather than incidence, and the limited temporal separation between behaviours and outcomes precludes causal inference. Finally, although we controlled for several individual-level sociodemographic variables associated with multimorbidity, we were unable to account for environmental and area-level factors that might contribute to differences in health outcomes [70].

### Practical implications and future research

By examining how behavioural risk factors cluster and are associated with disease, this study sheds light on how health behaviours and socioeconomic context jointly shape multimorbidity in UK adults. Further, our findings conceptually replicate those of Suhag et al. [19] in a broader age range, with similar patterns of behaviour stability and disease associations, strengthening confidence in these findings. Two clusters emerged as particularly vulnerable: *Overall Low-risk* and *Smokers*, both characterised by socioeconomic disadvantage and high disease burden. Conversely, clusters with multiple risky behaviours but favourable socioeconomic profiles had comparatively better health, again mirroring Suhag et al. [19]. Ethnic minority groups were over-represented in the cluster with the fewest risky behaviours (i.e. *Overall Low-risk)* yet did not have correspondingly better health outcomes [43, 54], underscoring the importance of structural causes of ill-health alongside individual behaviours.

Taken together, these findings suggest that clusters defined as ‘low risk’ on behavioural criteria alone may be systematically misclassified in risk stratification frameworks, with the potential consequence that prevention and screening efforts may overlook socioeconomically disadvantaged groups with high underlying disease burden. Moreover, the replication of these patterns across cohorts and age ranges further suggests that the observed behavioural–sociodemographic configurations are not cohort-specific, supporting their relevance for population-level prevention and intervention strategies that consider social context alongside behaviours. Future work should replicate these clusters using more comprehensive behavioural measures (particularly diet), examine disease incidence where possible, and link behavioural patterns to additional outcomes such as clinical biomarkers, disability, and service use to better identify vulnerable groups.

## Conclusion

We identified seven clusters each characterised by a distinct risk behaviour pattern, sociodemographic characteristics, and multimorbidity prevalence in a nationally-representative adult sample. The findings suggest that simply the number or combination of risk behaviours falls short in explaining the variations in disease outcomes across different clusters. Rather, some clusters defined by ostensibly low-risk behaviours were also characterised by socioeconomic disadvantage and therefore had counterintuitive associations with worse health outcomes. This underscores the role of social determinants in shaping health disparities and highlights the need to consider these factors in policy formulation.

## Declaration of Interests

We declare no competing interests.

## Contributors

JH, TLW, and AS conceptualised and designed the study. AS conducted the statistical analyses under the supervision of TLW and JH. AS wrote the first draft and all authors provided critical revisions and approved the final submitted version.

## Funding

AS was supported for this study by a studentship awarded to the Healthy Lifespan Institute and funded by the University of Sheffield. The funders had no say in the design and conduct of the study; collection, management, analysis, and interpretation of the data; preparation, review, or approval of the manuscript; and decision to submit the manuscript for publication.

## Ethical approval

The University of Essex Ethics Committee has approved data collection for the UK Household Longitudinal Study (UKHLS). Separate ethical approval and consent were not required for our secondary analyses as data were fully anonymised.

## Data sharing

The raw data involved in this analysis are available through the UK Data Service.

## Supporting Information

S1 Appendix. Supplementary Appendix File.

## Data Availability

The raw data involved in this analysis are available through the UK Data Service (https://www.understandingsociety.ac.uk/documentation/access-data/)

## Notes

### Competing Interest Statement

The authors have declared no competing interest.

